# Flattening the curve and the effect of atypical events on mitigation measures in Mexico: a modeling perspective

**DOI:** 10.1101/2020.05.21.20109678

**Authors:** M. Santana-Cibrian, M. A. Acuña-Zegarra, J. X. Velasco-Hernandez

## Abstract

On 23 and 30 March 2020 the Mexican Federal government implemented social distancing measures to mitigate the COVID-19 epidemic. We use a mathematical model to explore atypical transmission events within the confinement period, triggered by the timing and strength of short time perturbations of social distancing. We show that social distancing measures were successful in achieving a significant reduction of the effective contact rate in the early weeks of the intervention. However, “flattening the curve” had an undesirable effect, since the epidemic peak was delayed too far, almost to the government preset day for lifting restrictions (01 June 2020). If the peak indeed occurs in late May or early June, then the events of children’s day and mother’s day may either generate a later peak (worst case scenario), a long plateau with relatively constant but high incidence (middle case scenario) or the same peak date as in the original baseline epidemic curve, but with a post-peak interval of slower decay.

## 1 Introduction

On 23 March 2020, the Mexican Federal government officially established mitigation measures to control the COVID-19 epidemic. All public and private schools were closed and a set of non-mandatory social distancing recommendations was issued. One week later, on 30 March 2020, a Sanitary Emergency was declared, ordering the suspension of all non-essential activities in the public and private sectors, until 30 April 2020, a date that was later extended to 01 June 2020. The aim of these measures was to lower the incidence to manageable levels in terms of the expected number of critical cases [1]. On 16 April 2020, the federal government announced that, for Mexico City, the epidemic peak (day of maximum incidence) would occur on 08-10 May 2020 [2,3]. On May 8th, this estimate was corrected and moved to no later than 20 May 2020 [4,5]. A gradual lifting of social-distancing measures was announced to start on 01 June 2020. Unfortunately, government predictions seem to be underestimating the time of maximum incidence, since the epidemic curve has not shown clear signs of having reached the peak by mid-May. If conditions remain constant with respect to the population compliance of the mitigation measures, then the peak could occur by late May or early June [6,7], coinciding with the date set for the lifting of social-distancing. However, things may be more complicated than this. There are two holidays, children’s day and mother’s day, where mobility increases; these events may have an important impact on the epidemic curve and thus on the strategy to implement the lifting of social-distancing, not considered in the early forecasts.

In this paper, we explore the possible impact of such short time periods where the population does not follow the social distancing measures, and use Mexico City as an example to show the consequences of different scenarios. First, in Section 2 we explain the methods and data used for the analysis. Then, in Section 3, we describe the current situation of the COVID-19 pandemic in Mexico City, including an evaluation of the mitigation measures using the Richards model. Then, using a Kermack-McKendrick model [6], we generate scenarios that describe the impact of the two aforementioned holidays on the shape of the epidemic curve for Mexico City. Finally, the discussion and conclusions are presented in Section 4.

## 2 Methods

### 2.1 Data source

Data for the COVID-19 epidemic was provided by the Secretaría de Ciencia, Tecnología e Innovación of the Government of Mexico City through the COVID-19-CDMX database [8]. This data set contains details on all the confirmed and suspected COVID-19 individuals such as sex, age, residence region, date of symptoms onset, etc. Records are available from 22 February 2020 to 19 May 2020. Due to reporting delays and delays imposed by the incubation period of the virus, we do not use the last 14 days of data in all of our estimations.

### 2.2 Richards model

We fit the Richards model to the epidemic curve [9] to estimate the growth rate for two different periods, 22 February 2020 to 22 March 2020, and 23 March 2020 to 27 April 2020, to have an approximation of the reduction in the growth rate r resulting from the social distancing measures implemented by the federal government. We assume that the cumulative cases curve of COVID-19, *C*(*t*), can be described by the solution of

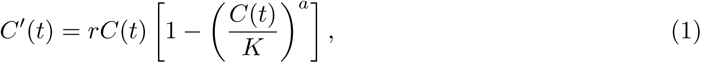

where *r* is the infection growth rate, *K* is the final epidemic size, and *a* is a scaling parameter to account for the asymmetry of the epidemic curve. This model is an extension of the simple logistic growth model that has already been used to predict cumulative COVID-19 cases in China [9].

The parameters *a*, *r*, and *K* must be estimated from the observed data. We use a statistical approach through Bayesian inference [10,11]. Technical details of the estimation process can be found in Appendix A.

### 2.3 Mathematical model setup

To evaluate the impact of atypical events, we use the model developed in [6] under the scenario that 70% of the general population is confined and 30% is not. This split of the population occurs when social distancing measures are implemented on 23 March 2020. The non-confined population includes workers with essential economic activities in government and industry, or individuals that work in the informal economy. In Mexico City, the informal economy represents 49% of the economically active population [12]. Once under confinement, this population abandons the confinement at a rate *ω* that we call the confinement-failure rate. This is the parameter we use as a proxy for population mobility. For more details on the mathematical model, we refer the reader to [6]. Figure 1 shows the model diagram.

**Figure 1:**
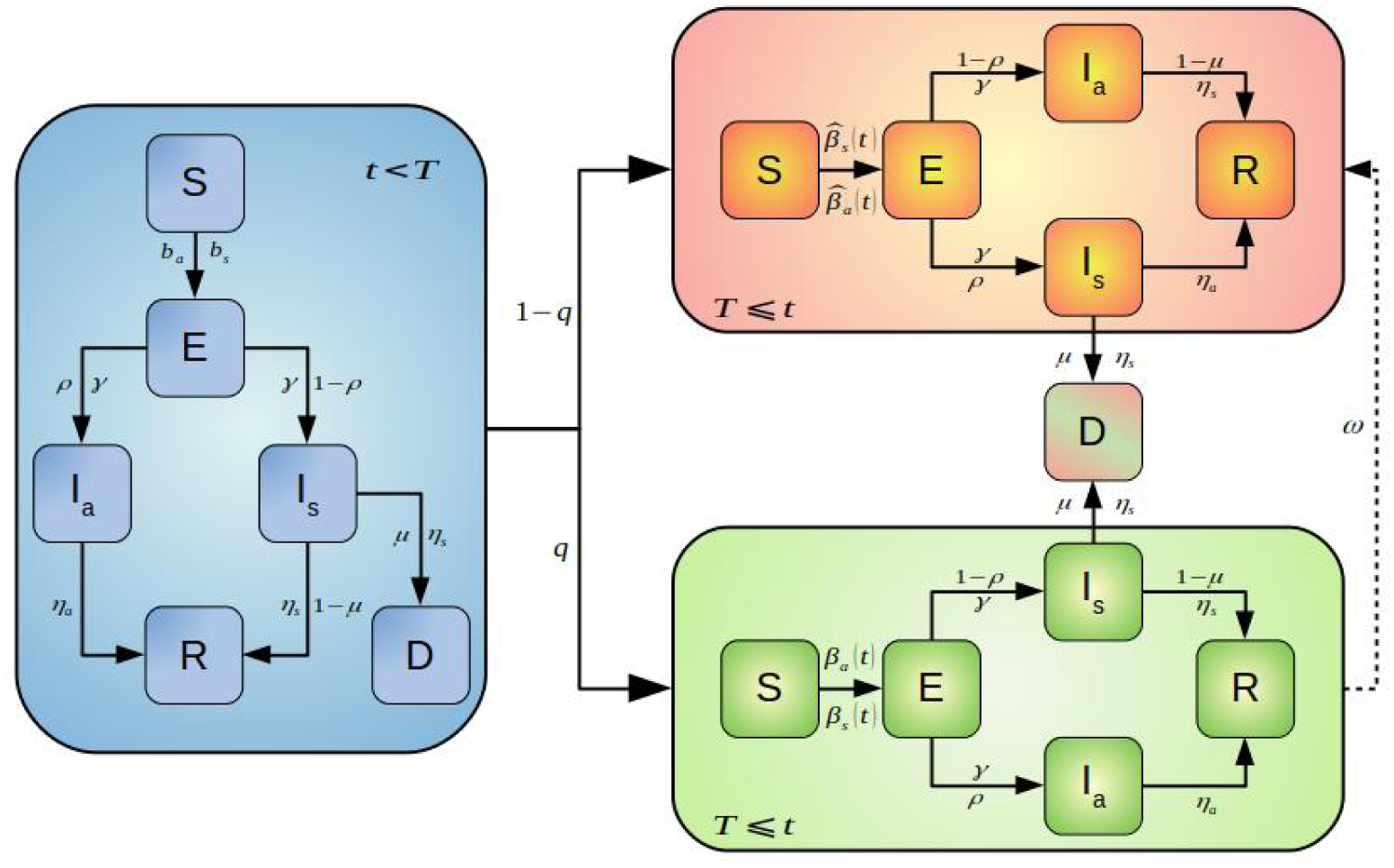
Structure of the mathematical model [6]. The state variables S, *E, I_a_, I_s_, R, D* represent the populations of susceptible, exposed, asymptomatically infected, symptomatically infected, recovered and dead individuals, respectively. Previous to the Sanitary Emergency measures the epidemics follows the dynamics represented in the blue diagram. Once Sanitary Emergency Measures are implemented the population splits into two: those who comply with the control measures (green box) and those who do not (pink box). The dashed line connecting the green and pink boxes represents the confinement failure rate *ω*.

Since individuals under confinement fail to comply with the social-distancing indicatives at a rate w (baseline *ω*_0_ = 0.005/day), they become part of the other subpopulation. The model assumes that, in both groups, the effective contact rate is actually reduced, but the subpopulation under confinement has the largest reduction of the two.

In the next section, we model the atypical increase in mobility on children’s and mother’s days as follows: i) in each case, we assume the increased mobility lasts only for a period of *τ* days, and ii) the increased mobility on these dates is reflected on an increase of the compliance-failure rate *ω*_0_ by a factor *k*. For these dates, the new compliance-failure rate is *kω*_0_.

## 3 Results

### 3.1 Effect of mitigation measures

Confirmed cases of COVID-19 epidemic in Mexico can be presented in several forms. Figure 2 shows confirmed cumulative cases reported by date of symptoms onset, by the date of arrival at the hospital, and by the date when tests were confirmed. It also shows that the growth pattern in each one is different. For example, if we focus on cases reported by the date of test confirmation (green bars), the cumulative cases show that the epidemic is still in the exponential growth period. On the other hand, for the cases reported by symptoms onset (blue bars), cumulative cases show linear growth. In this work we use this last representation (symptoms onset) since it describes the growth of the epidemic in terms of active cases. Figure 3 shows the number of confirmed and suspected cases by symptoms onset. Suspected cases are those individuals that present COVID-19-like symptoms but are still waiting for test results. As of 19 May 2020, in Mexico City, there are 6973 suspected cases, which amounts to 46% of the 15283 confirmed cases at the same date. Certainly, not all suspected cases will be confirmed as COVID-19 cases. If we consider that the positivity rate in Mexico City is approximately 35%, then 2440 of those 6973 suspects will be confirmed, a fraction that represents 16% of the total confirmed cases. The still increasing tendency of the number of suspected cases shown to date, lends support to the hypothesis that for Mexico City the peak of the epidemic has yet to be reached. This is an important consideration regarding the end of confinement measures and the impact of atypical events on the dynamics of transmission on the shape of the epidemic curve.

**Figure 2:**
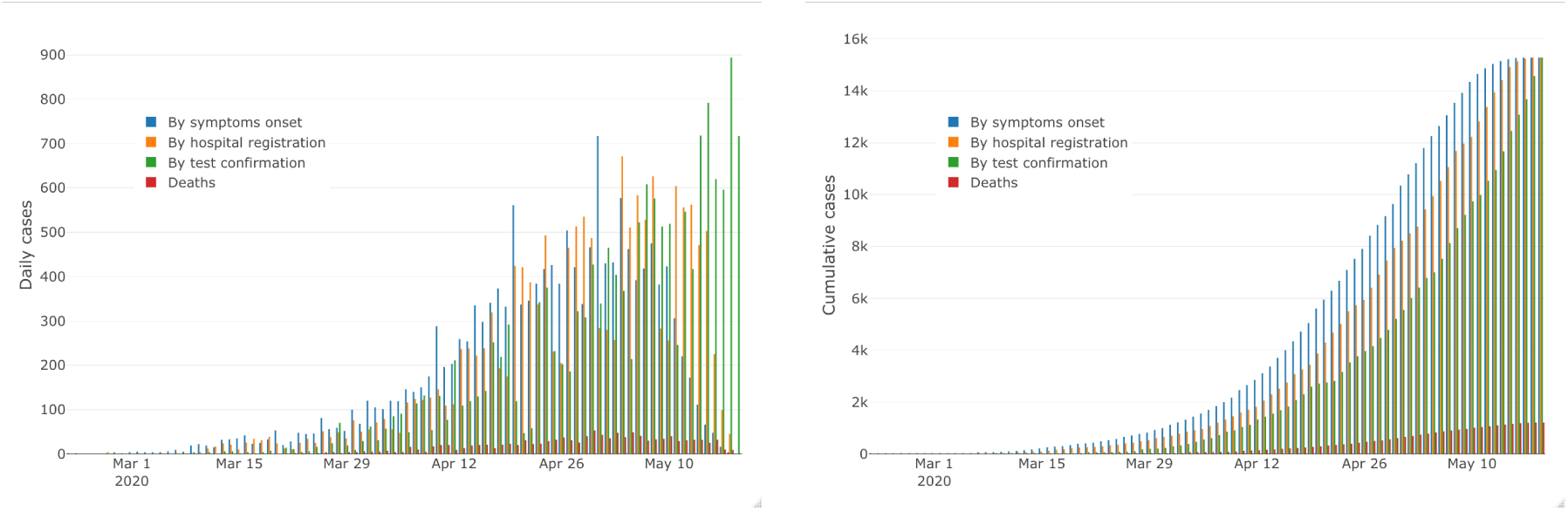
Daily and cumulative SARS-CoV-2 cases in Mexico City from 17 February 2020 to 19 May 2020 presented in three different forms: blue bars correspond to cases by symptoms onset, green bars are cases by date of arrival to hospitals, and yellow bars show cases by date when tests were confirmed. Deaths are also showed in red.

**Figure 3:**
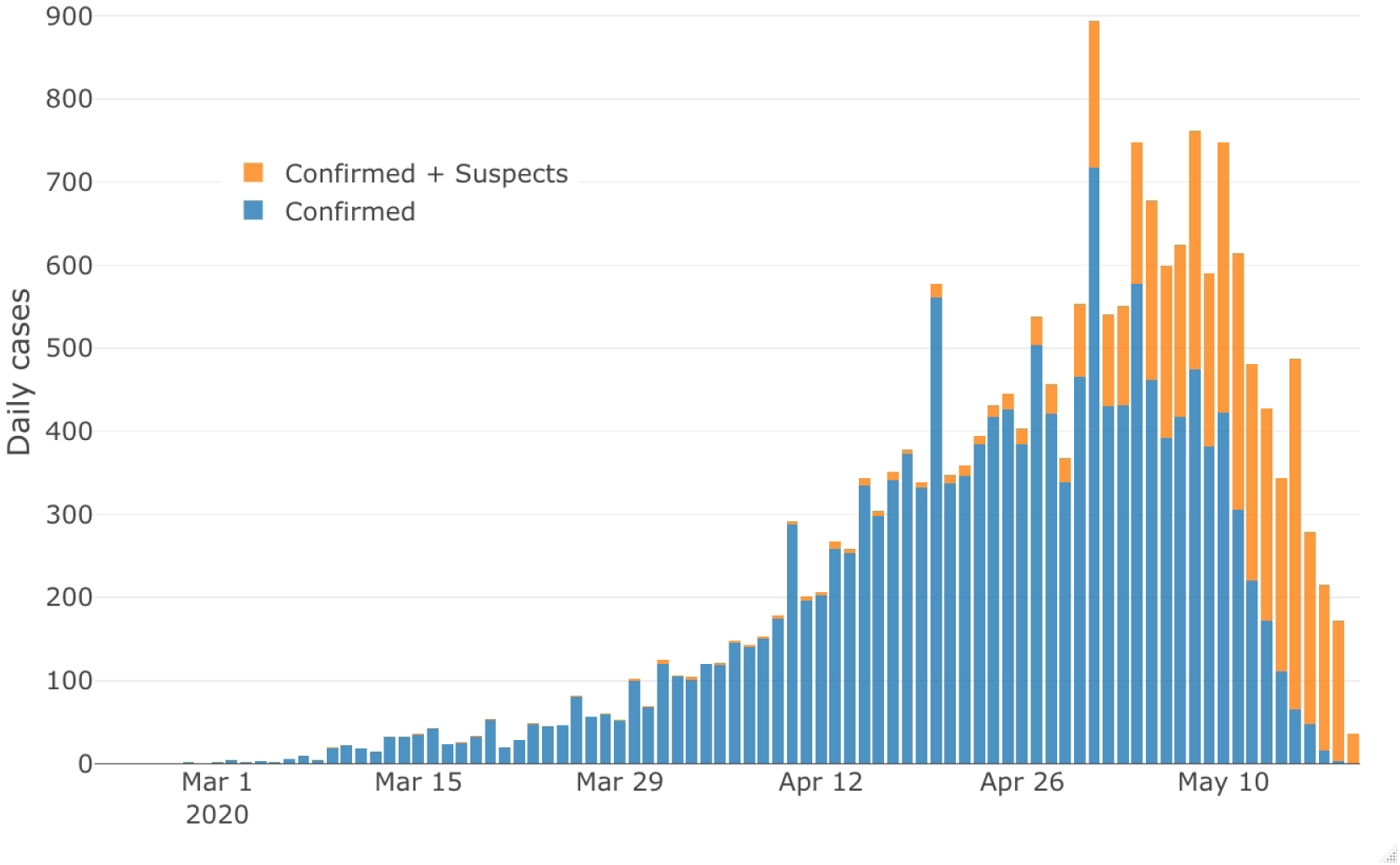
Daily confirmed and suspicious SARS-CoV-2 cases in Mexico City from 17 February 2020 to 19 May 2020. Blue bars correspond to cases by symptoms onset, orange bars correspond to confirmed cases plus the suspected cases.

We have already pointed out that some suspected cases are waiting for test results, but it is equally possible that many of them are yet to be tested. According to the official daily published data, the number of tests has dropped significantly since the first week of May (Figure 4). This does not necessarily mean that there are fewer tests but rather, that there is an important delay in the reporting of results. This possibility is consistent with Figure 3 that shows that the number of suspected cases has indeed increased since the beginning of May. It is possible too that there is a saturation in the laboratories that apply the tests and this could be causing a lag in the reported cases or that tests for the general population are being canalized elsewhere due to the increase in mortality and hospitals’ demand. This also could explain why there are many suspected cases that started symptoms in March and April that have not yet been confirmed. In summary, the most recent observations of the number of confirmed cases are still incomplete. It will take at least two weeks before we have data to conclusively decide whether Mexico City is really close to the maximum incidence peak or not.

**Figure 4:**
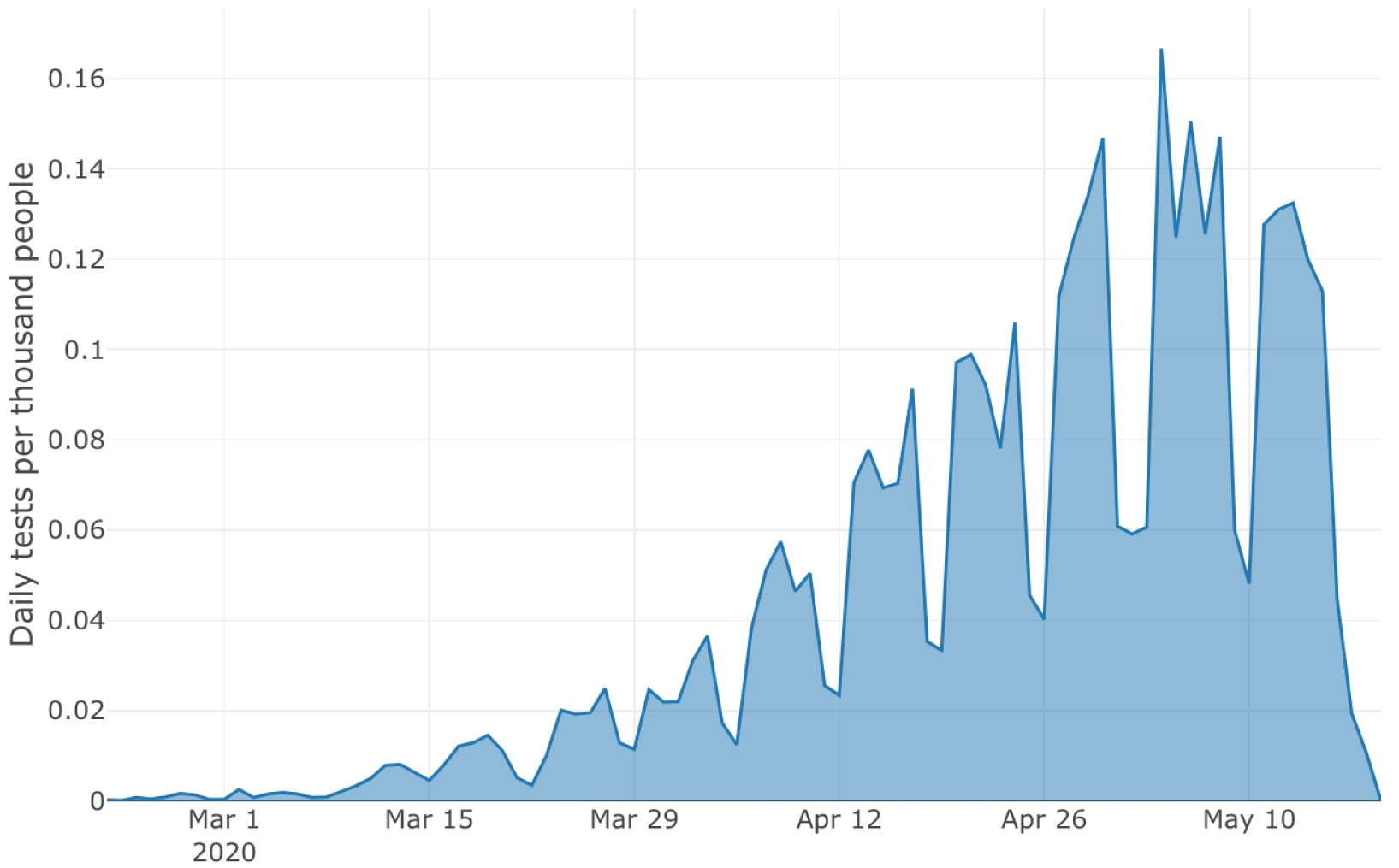
Total COVID-19 tests per thousand people in Mexico City from 22 February 2020 to 19 May 2020.

### 3.2 Reduction in epidemic growth rate

Table 1 shows the parameter estimates for the two periods. It can be seen that, before the start of social distancing on 23 March 2020, the growth rate is approximately 1.062 with a 95% interval (0.408,3.197). After 23 March 2020, the growth rate is 0.103 with a 95% interval of (0.096, 0.119). This gives us an average reduction of 90% of the epidemic growth rate in the early days of the implementation of social distancing measures. Figure 5 shows the observed daily data and the fit of the model before and after the isolation measures. Richards model also provides information about the maximum cumulative incidence (epidemic size). Our estimations project, at the date of writing, a maximum incidence to occur between 22-May and 15 June, 2020. Given the overall decreasing magnitude of the reproductive number *R_t_* (described below) at the time of writing, we cautiously provide results for *K* as an illustration of a possible scenario (see Table 1).

**Table 1:** Parameter median estimates and 95% posterior probability intervals before and after 23 March 2020. Here, *r* is the growth rate, *K* is the final size of the outbreak and *a* is a scaling factor.

|  | 22 February - 22 March |  |  | 23 March - 27 April |  |  |
| --- | --- | --- | --- | --- | --- | --- |
|  | Lower | Median | Upper | Lower | Median | Upper |
| $a$ | 0.009 | 0.031 | 0.148 | 0.296 | 0.510 | 0.698 |
| $K$ | 5,881 | 51,529 | 170,214 | 42,976 | 65,255 | 164,821 |
| $r$ | 0.408 | 1.062 | 3.197 | 0.096 | 0.103 | 0.119 |

**Figure 5:**
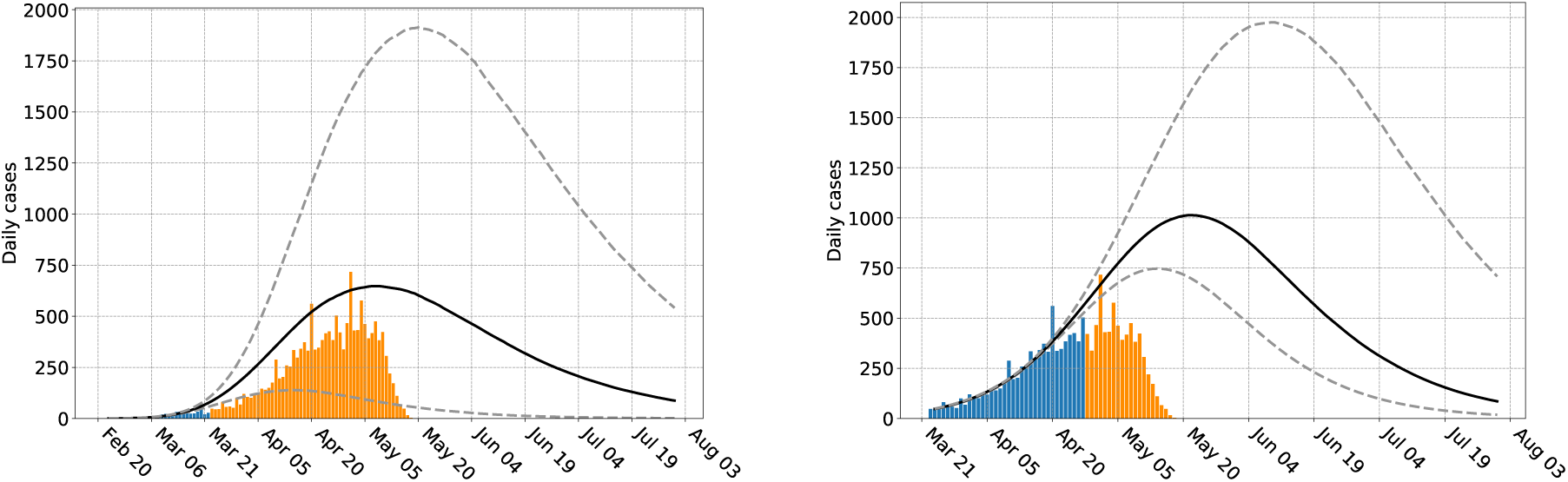
Daily observed cases and incidence obtained from Richards model: a) from 22 February 2020 to 22 March 2020, and b) from 23 March 2020 to 27 April 2020. Blue bars represent the observed data used to fit the model at each period, and orange bars represent the observed data that was not used. The estimates for maximum incidence are not used in this work.

Figure 6 shows the evolution of the instantaneous reproduction number *R_t_* in Mexico City until 5 May 2020, four days after mother’s day. It is computed using the algorithm in [13,14] with a mean intergenerational period of 4.7 days [15]. Note that the *R_t_* trend shows a slight increase just around 30 April and 01 May. Transmission events occurring in these days will be reflected within 14 days, both as increased incidence and mortality. At the date of writing, we still do not have the information to explore this possibility. We can conclude, however, that although the social-distancing measures were indeed effective by reducing transmission as evidenced by the reduction of the epidemic growth rate after 23 March 2020, they also pushed the peak towards the end of the month, at the earliest.

**Figure 6:**
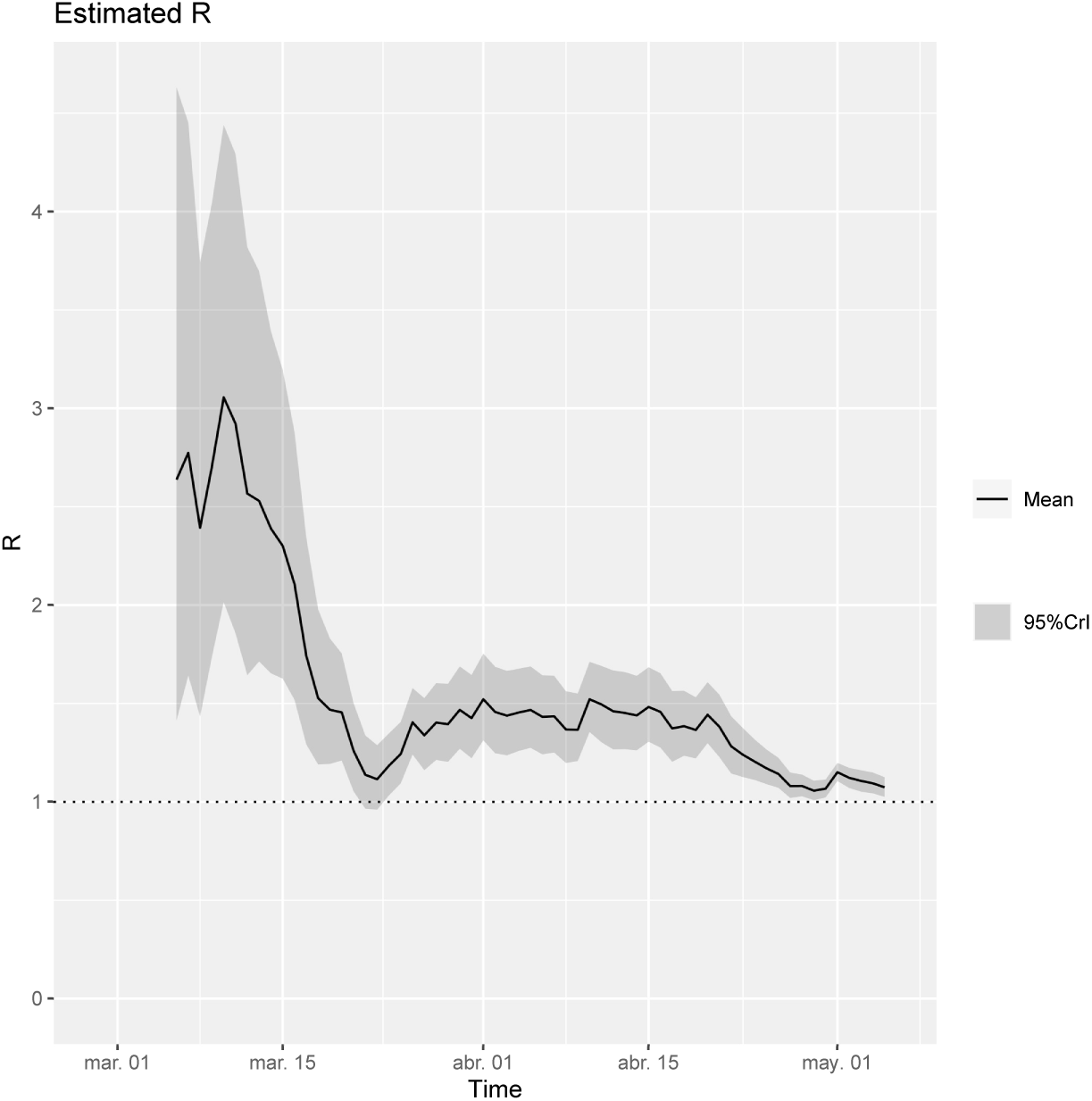
Instantaneous reproduction number for Mexico City using a median serial interval of 4.7 days following the study in [15]. The Figure shows the estimates from 28 February 2020 to 05 May 2020. A clear jump can be observed for 30 April and 01 May, indicating a likely increase in active transmission during these days.

### 3.3 Effect of atypical events on social-distancing measures

In Mexico, social-distancing measures are focused on social-distancing and non-pharmaceutical interventions (NPIs) to reduce contact between individuals. However, in Mexico City, increases in population mobility that last a few days have been observed [16]. This increased mobility weakens the strength of the NPIs and, therefore, may have an impact on disease transmission. This section explores the consequences of bursts of increasing mobility near the peak day of the epidemic curve.

As already mentioned before, there are two important holidays (in terms of population mobility) within the period of confinement: 30 April, children’s day, and 10 May, mother’s day. There exists evidence that population mobility slightly increased [16] in these two days. To study the effect, we use the periods A): 29-30 April 2020, which are weekdays and B): 08-10 May 2020, a weekend. For these periods, the confinement-failure rate is *kω*_0_, where *k* = 2 or 5. The magnitude of k is arbitrary but, relative to *ω*_0_ = 0.005/day, it is a small perturbation.

Our scenarios are:

- Scenario I with two cases: I.1) *k* = 2 in A and B, I.2) *k* = 5 in A and B.
- Scenario II with two cases II.1) *k* = 2 in A, *k* = 5 in B; I.2) *k* = 5 in A, *k* = 2 in B.

Scenario I assumes that both periods have equal *ω*. This is a baseline case since mother’s day in Mexico is an extremely important family date whose popularity is well beyond that for children’s day. Scenario II addresses this difference. It considers an unequal outflow from social-distancing. We look at the impact of these two events on the epidemic peak, assuming that the peak occurs in early June.

Figure 7 shows our simulations for both scenarios. Case I.2 is clearly the worst-case scenario (black dashed line). After the perturbation, the incidence peak is higher and later for several weeks. The epidemic size increases too. Scenario I.1 (black line), on the other hand, is comparatively benign. The peak is still reached on the baseline date, then there is an interval of about a month where the epidemic curve decreases with a lower rate than the baseline case. Afterward, the decrease is essentially at the same speed as that for the original curve (blue). Case II.1 (red dashed line) occurs when the children’s day period (29-30 April) has a lower confinement-failure rate than the mother’s day period (08-10 May). Observe that when the peak is reached, the incidence curve does not show a significant decay but rather, enters into a plateau phase that lasts several weeks after the baseline peak date (blue curve). On the other hand, in scenario II.2 (red line), the largest increase in the confinement-failure rate occurs for fewer days (children’s day period), producing an epidemic curve similar to the one in scenario II.1.

**Figure 7:**
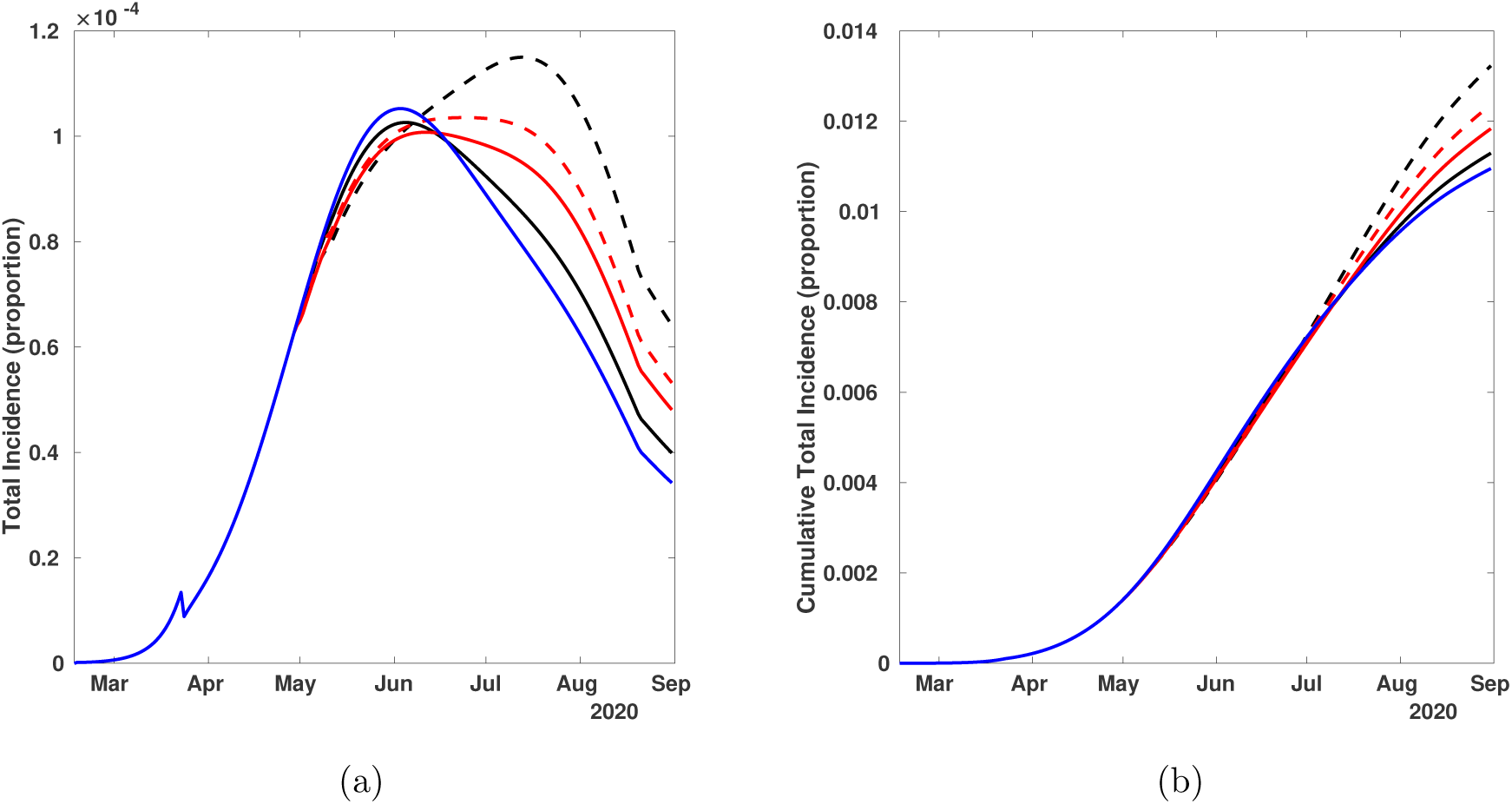
Impact of two atypical events of high mobility, occurring on 30 April 2020 and 10 May 2020, on the baseline epidemic curve (blue line) with peak incidence on 03 June 2020. Scenarios I.1, I.2, II.1, and II.2 are represented by black line, black dashed line, red dashed line, and red line, respectively. a) daily incidence, b) cumulative incidence. In this plot, the increase of the total number of cases produced in the different scenarios is evident.

#### Remark

We illustrate the effect of these atypical events with the specific holidays above, but the exercise can be viewed in more general terms. Figure 8 shows the consequences of increased mobility in short periods occurring before or after the incidence peak. In this case, the mobility increases for three days by a factor of *k* = 10 times the baseline containment failure rate (i.e., *ω* = 10*ω*_0_). The worst case scenario occurs when the atypical event is located on the exponential growth phase of the curve and far from the maximum incidence (black discontinuous line). A less extreme scenario occurs when the increase in population mobility is located after and far from the incidence peak (green discontinuous line). For completeness, in Appendix B, we show simulations for a mobility increase of only five-fold the baseline value. This scenario is, hopefully, the one that will occur in Mexico City, but at the time of writing, the incidence data for the days around 10 May 2020 is still incomplete and the mobility impact cannot be yet verified.

**Figure 8:**
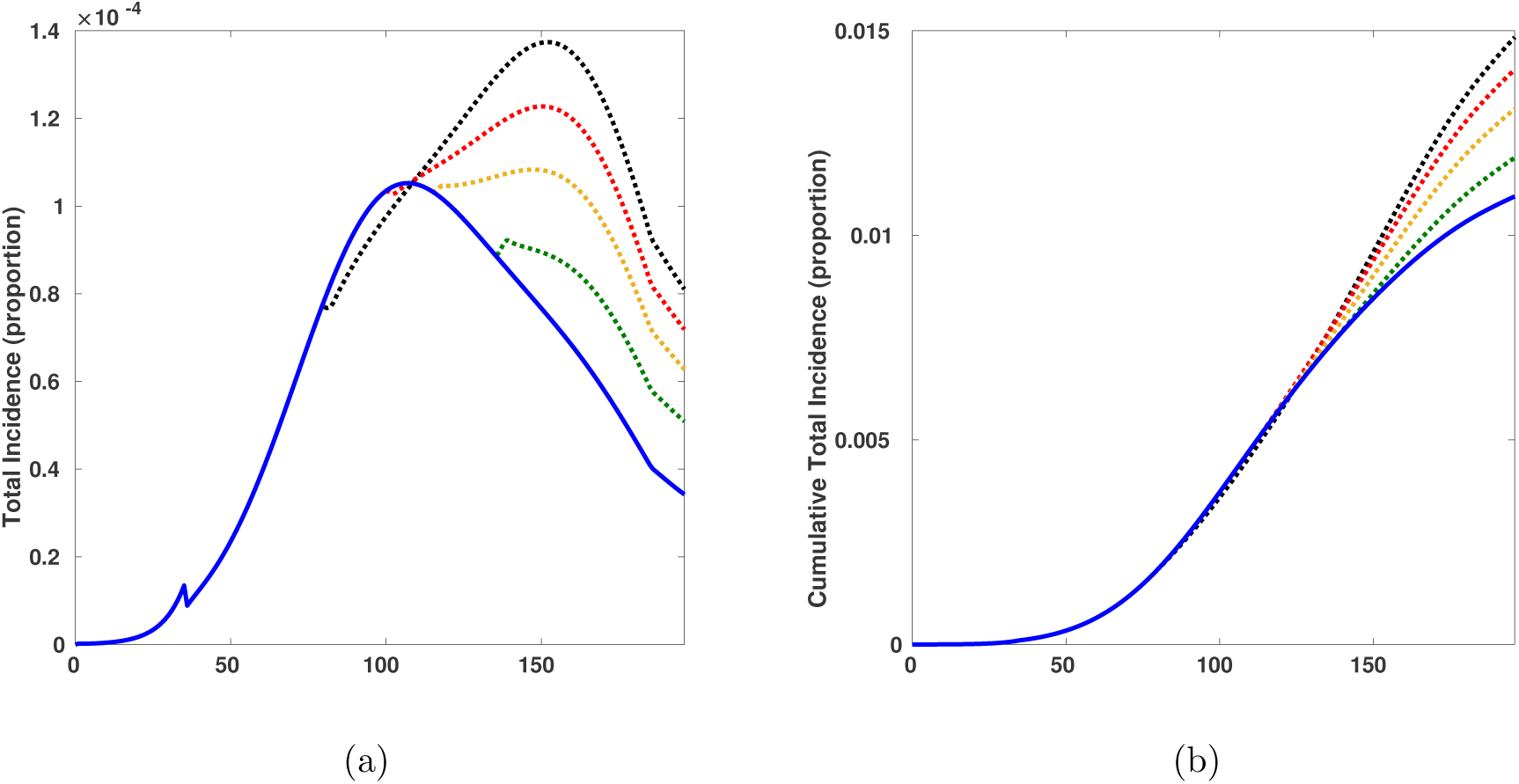
Impact of different times of mobility increase on the epidemic curve. Mobility increase is, for all cases, 10*ω*_0_. Blue line, baseline epidemic curve. Black, red, gold, and green discontinuous lines show the scenarios when the mobility event starts four weeks before, a week before, a week after, and four weeks after peak incidence, respectively.

## 4 Conclusions

We have presented an analysis of the current COVID-19 epidemic in Mexico, illustrating some of its features with the case of Mexico City. Our analysis shows that social distancing measures implemented by the country’s federal health authorities on 23, 30 March 2020 were effective in mitigating the epidemic growth rate in the weeks after this date. This result is supported by our estimate of the reduction of the epidemic growth rate using Richard’s model, and also with the sustained downward trend of the effective reproductive number until 29 April 2020 (right up to children‘s day, on 30 April). The analysis of the precise epidemiological situation in Mexico is difficult because, among other things, the COVID-19 test rate is very low [17]. Moreover, its high positivity rate (in the interval 21% to 40% in Mexico City for the first week of May [8]) indicates a large under-reporting of cases that affects the estimates of true mortality and incidence rates. Our analysis relies on the confirmed cases reported by the General Directorate of Epidemiology. Nevertheless, the available data shows that although the aim of flattening the epidemic curve has been achieved in Mexico City and, on average, in the whole country, the Mexican case shows that mitigation measures cannot be only concerned with spreading the infections over a longer period and reducing the incidence peak. Timing is of utmost importance [18]. For Mexico City, the government originally forecasted the peak to occur by 8-10 May 2020, setting the date for liberation of mitigation measures for 30 May. The peak has not yet been reached by 20 May (when this report is being written), pushing the likely dates for it to occur either close to the end of May or, likely, beyond 01 June 2020, the new revised date set for lifting mobility restrictions and the start of the reactivation of the economy. In this context, events where a high increase in mobility takes place, like what possibly occurred on children’s day and mother’s day (30 April and 10 May, respectively), may impact the epidemic curve before the peak occurs.

We have used a mathematical model previously developed [6], to generate plausible scenarios that may affect the epidemic curve, as related to the timing and strength of perturbations of social distancing measures. We have explored the effect of pulses of unusual activity (within the confinement period) on the epidemic curve of COVID-19 in Mexico City. These are necessarily theoretical results, but we believe they illuminate the importance of counting with reasonable estimates for the timing of maximum incidence. We have already mentioned that mitigation measures will be lifted on 01 June 2020 as announced in early April 2020, and that the current epidemic trend strongly indicates that the peak will occur past the middle of May. If it was to occur in late May or early June as predicted by other models [6,7], then the events of children’s day and mother’s day may either generate, depending of the magnitude of *ω*, a later peak (worst case scenario), a long plateau with relatively constant but high incidence (middle case scenario) or the same peak date as in the original baseline epidemic curve but with a post-peak interval of slower decay (Figures 7 and 8).

Mathematical models are essential in the fight against COVID-19. They are tools for evaluating mitigation measures, estimating mortality and incidence, and projecting scenarios to help public health decision-makers in their very difficult and important task of controlling the epidemic. In this paper, we have used mathematical models to evaluate and generate scenarios. Although precise forecasting is not our aim, we consider that these results can be helpful for decision-makers.

## Data Availability

Data is publicly available.

https://datos.cdmx.gob.mx/explore/dataset/base-covid-sinave/table/

## Authors Contributions

All authors contributed equally.

## Acknowledgments

JXVH acknowledges support from DGAPA-PAPIIT-UNAM grant IN115720. We thank the advice and encouragement of Dr. Héctor Benitez, Dr. Ramsés Mena, and Dr. William Lee Ardavín from UNAM. We also thank Ruth Corona for their help during the preparation of this manuscript, and the Secretaría de Ciencia, Tecnología e Innovacíon of the Government of Mexico City for facilitating the data on the COVID-19 epidemic.

## A Parameter estimation of the growth rate

Let *Y_j_*, for *j* = 1, 2, …, *n*, be the number of observed cumulative cases at time *t_j_*, with *t_j_* given in days. We assume that *Y_j_* follows a Negative Binomial distribution with mean value *C*(*t_j_* |*a,r,K*) and dispersion parameter *α*. Here, *C*(*t_j_* |*a,r,K*) is the solution of Richards model presented in (1). Assuming that, given the parameters, the observations *Y*_1_, *Y*_2_, …, *Y_n_* are conditionally independent, then

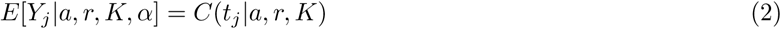

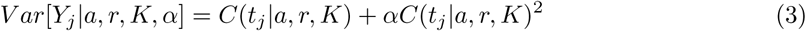

The Negative Binomial distribution allows to control the variability of the data by considering over-dispersion, which is common for epidemiological data. If *α* = 0, then we return to the Poisson model which is often used in this context.

Let ***θ*** = (*a*, *r*, *K*, *α*) be the vector of parameters to estimate. The inclusion of the parameter *α*, which is related to the variability of the data, not to the Richards model, is necessary since in practice this variability is unknown. Then, the likelihood function, which represents how likely it is to observe the data under the Negative Binomial assumption and Richards model if we knew the parameters, is given by

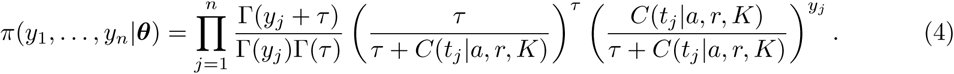

Consider the parameters *a, r, K*, and *α* as random variables. Assuming prior independence, the joint prior distribution for vector ***θ*** is

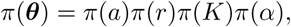

where *π*(*a*) is the probability density function (pdf) of a Uniform(0,1) distribution, *π*(*r*) is the pdf of a Uniform(0,5), *π*(*K*) is the pdf of a Uniform(*K*_min_, *K*_max_), and *π*(*α*) is the pdf of a Gamma(shape=2, scale=0.1). To select the prior for parameter *r*, we consider that other estimations of *r* are close to 0.3 [9]. In addition, there is no available prior information regarding the final size of the outbreak *K*. This is a critical parameter in the model and, to avoid bias, we assume a uniform prior over *K*_min_ and *K*_max_. To set these last to values, we consider that the minimum number of confirmed cases is the current number of observed cases *Y*(*t_n_*) times 10 and 5, i.e., *K*_min_ = *y_n_* * 10 and *K*_min_ = *y_n_* * 5, for the periods before and after 23 March 2020, respectively. To set the upper bound for *K*, we consider a fraction of the total population *K*_max_ = *N* * 0.02, where *N* is the population size of Mexico City. This fraction was determined base on the observations of other cities such as New York, where the total population size is similar to Mexico City and the proportion of infected represents one of the worst case scenarios up to date.

Then, the posterior distribution of the parameters of interest is

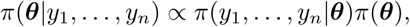

and it does not have an analytical form because the likelihood function depends on the solution of the Richards model, which is non-linear in the parameters. We analyze the posterior distribution using an MCMC algorithm that does not require tuning called *t-walk* [19]. This algorithm generates samples from the posterior distribution that can be used to estimate marginal posterior densities, mean, variance, quantiles, etc. We refer the reader to [10] for more details on MCMC methods and to [11] for an introduction to Bayesian inference with differential equations.

## B Low rate of mobility

Figure 9a shows the change in the epidemic curve when mobility increases for a single period lasting three days. We show how the location of this perturbation with respect to the peak date, affects the epidemic curve for a low confinement-failure rate *ω*. If the perturbation occurs in the exponential growth phase, the total incidence (area under the curve Figure 9b) will be higher than when the event is located further to the left of the peak. When the event occurs in the declining phase of the epidemic outbreak, the incidence continues decaying but at a slower pace, generating a net increase on the total number of cases.

**Figure 9:**
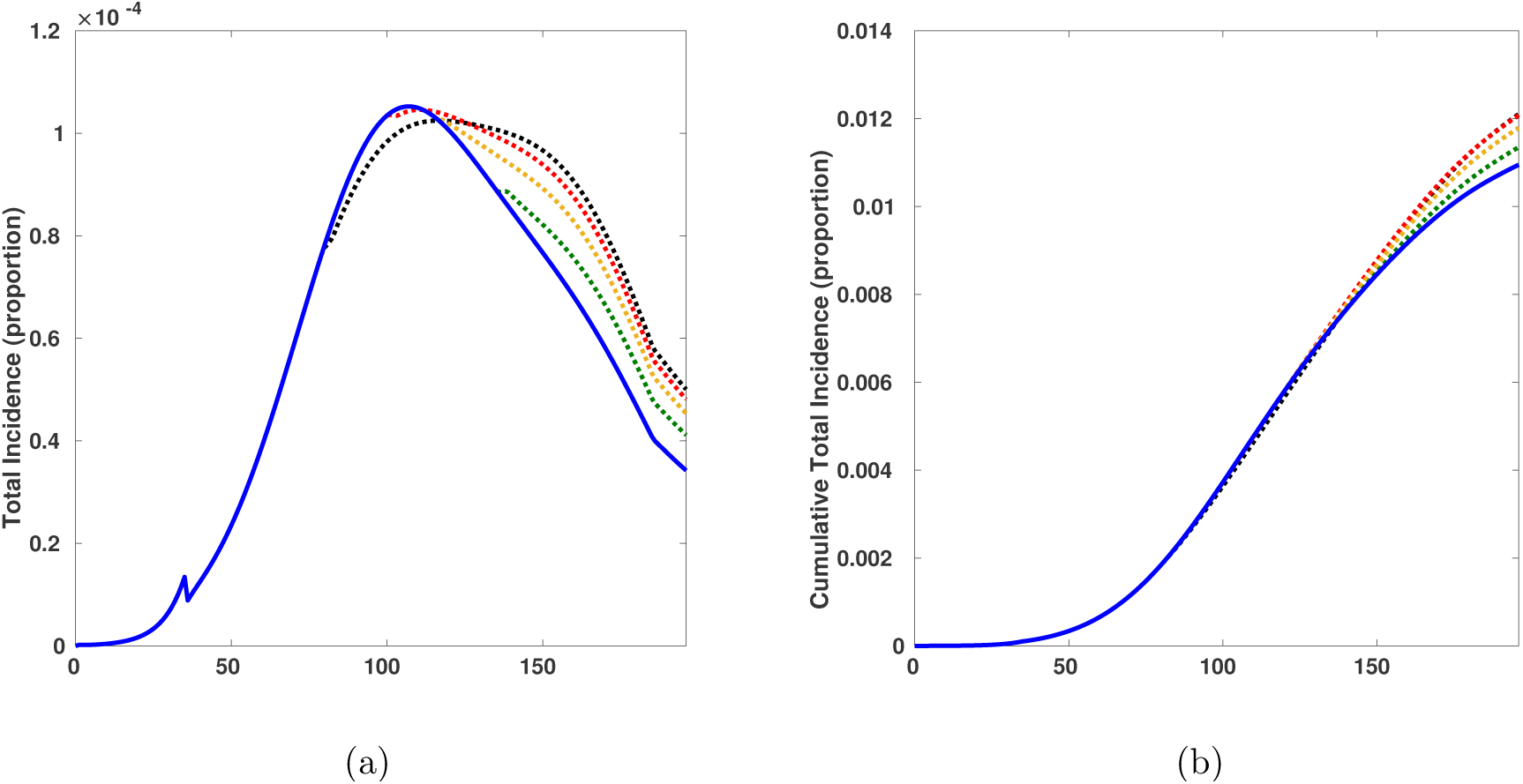
Impact of different times of mobility increase on the epidemic curve. Mobility increase is, for all cases, 5*ω*_0_. Blue line, baseline epidemic curve. Black, red, gold, and green discontinuous lines show the scenarios when the mobility event starts four weeks before, a week before, a week after, and four weeks after peak incidence, respectively. a) Daily incidence curve; b) cumulative incidence curve. The equivalent effect for a May 10-like type of scenario on the curve, is represented by the black dashed line in both panels. In b) is coincides with the red dashed line.

